# Integrity performance assessment of a Closed System Transfer Device syringe adaptor as a terminal closure for Luer-Lock syringes

**DOI:** 10.1101/2021.11.11.21266172

**Authors:** Kate E Walker, Romana Machníková, Laima Ozolina, Alan-Shaun Wilkinson, Andrew J Johnson, Navneet Bhogal, Kate Pegg

## Abstract

**Objectives:** To investigate the container closure integrity of a Closed System Transfer Device syringe adaptor lock in combination with disposable Luer-Lock syringes as the terminal closure device. The United Kingdom (UK) National Health Service (NHS) Pharmaceutical Quality Assurance Committee (PQAC) requires syringe integrity data for final storage devices of aseptic products such as chemotherapy drugs when prepared in advance and stored prior to use as is standard practice for dose banded drugs. The assessment comprised both physical and microbial integrity testing of the combination closed system/ Luer-Lock syringes containers at syringe sizes: 1mL, 20mL, and 50mL.

**Methods:** Integrity testing was performed as described in the NHS Pharmaceutical Quality Assurance Committee Yellow Cover Document 2^nd^ edition 2013 “Protocols for the Integrity Testing of Syringes” with Chemfort™ (Simplivia, IL) syringe adaptor lock (SAL) devices as replacement for sterile blind hubs. Microbiological integrity was assessed according to Method 1 part 1.4 using *Brevundimonas diminuta* at 32°C for up to 14-days contact time. Two positive control devices per syringe size were tested using a blind hub cap as closure which was loosened prior to test. Physical integrity was assessed using Method 3 of the yellow cover document which is a dye intrusion method. Dye intrusion was assessed both visually and using a validated ultraviolet-visible spectrophotometer method. For each size/ batch of test articles a positive control device (n=1) was assessed using a wire wrapped around the syringe plunger tip deliberately compromising integrity. Negative controls for each size (n=1) consisted of devices not immersed in methylene blue dye.

**Results:** Chemfort™ syringe adaptor lock/ Luer-Lock syringe combinations were shown to be: (1) free of microbiological contamination after 14-days contact time (n=60), (2) free of dye intrusion at all syringe sizes tested (n=61 in total). The data demonstrates 100% closure integrity of the final container system when Chemfort™ syringe adaptor lock replaces the syringe hub as the terminal closure device. All positive control devices demonstrated system suitability as container integrity was compromised in all positive control tests. All negative controls were negative for microbial and dye intrusion.

**Conclusions:** Syringe adaptor lock components complied with the NHS Pharmaceutical Quality Assurance Committee Yellow Cover Document syringe integrity requirements when used as the terminal closure of Luer-Lock disposable syringes from 1mL up to 50mL. Therefore, syringe adaptor lock (Chemfort™) can be used as the terminal closure system for pre-filled syringes of chemotherapeutic drug products prepared in advance in UK NHS Pharmaceutical Technical Services.

**What this paper adds:** *What is already known on this subject:*

- There is a paucity of data to support the use of closed system transfer device (CSTD) components as terminal closure devices in combination with single use leur-lock syringes in pharmaceutical technical services in the UK.
- A recent publication in 2020 by Marler-Hausen *et al*. demonstrated reduction in contamination on a UK hospital day ward when a Chemfort™ syringe adaptor CSTD components were used, providing better protection for healthworkers.
- National Health Service (NHS) Quality Assurance Committee Yellow Cover Document (YCD) for “Protocols for the Integrity Testing of Syringes” states container integrity data is required to support use of CSTD syringe caps in aseptic services and enabling their greater use in administration.

*What this study adds:*

- This study provides the necessary integrity data for Chemfort™ (Simplivia, IL) syringe adaptor lock components in combination with syringe. Physical and microbiological integrity complied with NHS YCD requirements. Chemfort™ is the first CSTD to meet the YCD acceptance criteria allowing components to be used in aseptic services including Dose-Banding in the UK.

## INTRODUCTION

Hazardous drugs (HDs) such as antineoplastics are routinely prepared in hospital pharmacy and used in the treatment of patients suffering from various forms of cancer, the scale of which is increasing due to longer life expectancy amongst the population worldwide.^1,2^ However, whilst there is a defined benefit to the patient, accidental exposure of the healthcare workers to the same HDs can result in harm with no associated benefits.^3,4^

Anecdotal evidence suggests that health workers are becoming harmed by accidental exposure to HD materials and this is the subject of a number of papers and reviews, with the risk of exposure to healthcare workers prompting many countries to develop guidelines for safer handling of hazardous drugs.^4-6^ One intervention that has the potential to reduce unintended occupational exposure to HDs is the use of closed system transfer devices (CSTD’s).^7-9^

Closed system transfer device (CSTD) components are designed to allow safe transfer of hazardous drug materials, thereby minimising healthcare worker exposure both during drug preparation and administration. CSTD’s are defined by the National Institute for Occupational Safety and Health (NIOSH) as “a drug transfer device that mechanically prohibits the transfer of environmental contaminants into the system and the escape of hazardous drug or vapor concentrations outside the system”.^3^ Two CSTD technologies exist in the market, and both prevent accidental release of aerosols, vapour and liquids: (1) physical barrier and (2) air filtration technology.^10,11^ Recently USP<800> was introduced which is the only pharmacopeia that mandates for CSTDs to be used in hazardous drug administration and recommends their use for drug preparation.^12^ In the UK, the NHS Pharmaceutical Quality Assurance (NHSPQA) committee has recently published guidance advocating the use of CSTD syringe components as replacement of the storage cap immediately prior to connection to the patient for cytotoxic chemotherapy administration.^13^ The guidance document also states that IV bags should be of the needle free variety or be spiked with closed system devices for safe handling of cytotoxic drugs in clinical areas. ^13^ The NHSPQA guidelines specifically requests manufacturers of CSTDs to provide necessary device integrity data along with product compatibility and stability data to “enable closed system syringe caps to be able to be added in aseptic services”.^13^ Adding CSTD components as part of the final closure system for pre-filled syringes in pharmacy technical services (PTS) reduces the potential for accidental exposure to nursing staff when removing the syringe cap prior to administration as this represents an “open” system. A recent study by Marler-Hausen *et al*. reports a significant reduction in exposure when syringe caps are replaced with CSTD components on the ward prior to administration compared with no CSTD.^9^ The current UK guidance where syringe caps are replaced by CSTD components on the ward prior to administration involves “opening” the system and therefore creates potential for exposure.^13^ Therefore there is an urgent operational need for device integrity data relating to CSTD components when used in combination with syringes to enable CSTD components to be added in PTS during compounding rather than at ward level.

In the UK, the gold standard for testing of syringe integrity is the NHSPQA yellow cover guidance document (YCD).^14^ The YCD requires that both microbiological and physical integrity tests are performed on the container closure system to assess integrity. Microbiological sterility testing of media fills (n=20) must be performed following immersion of the container in a culture broth according to either method 1 or 2 of the YCD.^14^ Method 2 has a short contact time of 30 minutes and employs *Escherichia coli* as the challenge agent. In the present study method 1 was selected which has an extended contact time of 14-days at temperature of 30-35°C and uses the challenge organism *Brevundimonas diminuta* which provides for a more stringent test of integrity.^14^ The test for physical integrity (Method 3) of the closure device is a dye intrusion test using methylene blue dye with the devices rotated for 2 hours whilst immersed in a solution of the dye.^14^ The physicochemical and microbial tests described in the NHS YCD share significant commonality with other methods described within USP <1207> and European Pharmacopeia (Ph. Eur. 3.2.9) for container closure integrity (CCI) testing.^15-17^ Although there is increasing preference for deterministic methods to be used in container closure integrity testing dye intrusion testing which is a probabilistic method remains one of the most commonly applied methods.

For adoption of CSTD components in PTS in the UK, syringe adaptor components must be tested to meet the NHS YCD standards for syringe integrity testing, whereby the syringe adaptor component replaces the sterile blind hub as the terminal closure.^14^ The present study aims to assess container closure integrity of a Closed System Transfer Device syringe adaptor lock (SAL) in combination with disposable Luer-Lock syringes as the terminal closure device.^13,18-19^ The present study has the potential to provide the necessary data to support adoption of SAL closed system device components within PTS as replacement for syringe hubs where there is currently a paucity of data.

A Cochrane review was recently published by Gurusamy *et al*. which concluded that there was no evidence to support or refute the routine use of closed-system drug transfer devices in addition to safe handling of infusional hazardous drugs over safe handling on its own.^20^ The conclusion was based on the low quality evidence of differences in healthworker exposure between CSTD plus safe handling versus safe handling alone.^20^ Since the initial Cochrane review was published a number of criticisms including those made in the American Journal of Health-System Pharmacy with commentary by McDiarmid *et al*. have been published along with rebuttal articles by various researchers working in the field of occupational health and the oncology community.^21-23^ A full scientific discussion has also been published in a rapid letter format by Gurusamy *et al*. also in the American Journal of Health-System Pharmacy addressing the criticism levelled at the review by McDiarmid and colleagues.^24^ This resulted in a revised Cochrane review published with adjusted findings not dissimilar to the original conclusions.^25^ The debate continues however and McDiarmid *et al*. has since published a follow up article stating that the criticisms of the Cochrane review remain valid.^26^ This discussion is outside of the scope of the present study.

Despite some concerns over the quality of evidence supporting CSTD use, the 2018 NHS yellow cover document “Guidance on handling cytotoxics in clinical areas” states that “Closed system caps should be added to syringes for IV use following removal of the storage cap immediately before connecting to the patient”.^13^ The same NHS guidance document also states that “CSTD device manufacturers/ suppliers should provide the required information in terms of integrity and product compatibility and stability to enable closed system syringe caps to be able to be added in aseptic services” – this study aims to provide the much needed data to support with the first of these requirements relating to CSTD use in aseptic services.^13^

## MATERIALS AND METHODS

### Materials

Chemfort™ Syringe Adaptor Lock (SAL) which is a CE marked CSTD component (MG245277, Simplivia Healthcare ltd, Kiryat Shmona, IL) were used as the tested CSTD components in this study. Chemfort™ Vial Adaptor (VA) (MG245248, Simplivia Healthcare ltd, Kiryat Shmona, IL) were used to allow draw up of media. All manipulations of the Chemfort™ SAL and VA were performed in accordance with the manufacturer’s instructions for use (IFU).^19^ Becton Dickinson (BD) disposable Luer Lock (LL) syringes of sizes: 1mL (10630694, Fisher UK), 20mL (10569215, Fisher UK) and 50mL (10636531, Fisher UK) were assessed as drug containers. Single strength tryptic soya broth (TSB) (Cherwell Laboratories, Bicester, UK) was used as the growth media. Sterile blind hubs were used for control devices (BD, UK). *Brevundimonas diminuta* (ATCC 11568) was supplied by LGC Standards in the UK. TSA 90 mm plates (IRR Cherwell, UK) and TSA + Neutraliser number 4 60mm plates (IRR Cherwell, UK) were used for growth of cultures. Prochlor 8-hour sporicidal wipes (Contec, UK), IMS 70% Alcohol wipes (Helepet, UK), sterile wipes (individually wrapped) (Helapet, UK) and IMS 70% Ethanol spray (Helapet, UK) were used for disinfection. 3,7-bis(Dimethylamino)-phenothiazin-5-ium chloride (methylene blue) dye (0.4%) CAS 61-73-4 (Sigma Aldrich, UK) was used for dye intrusion testing. MilliQ >18 Mega Ohm purified water was generated prior to use (Elix Merck Millipore, UK). Small split cotter pins and wood screws were used to secure the syringe plungers (Machine Mart, UK) and Leifheit storage containers were used for immersion of devices (Amazon, UK) for dye intrusion testing. Uv-vis grade flat bottomed 96 microwell plates were used for absorbance reading all solutions from dye intrusion tests (Fisher, UK).

### Equipment

Spectrometer Epoch plate reader (Biotek, UK) was used to measure absorbances at 660nm. Roller mixer (Stuart SRT9D, Fisher, UK) was used for dye intrusion tests. Validated grade A horizontal laminar flow cabinet (HLF6B, Envair UK) used for device preparation. LEC incubators (300WNP, LEC UK) and (300NP, LEC UK) used for incubation of all cultures and were identical in operation. All incubators were monitored continously using the COMARK temperature monitoring system.

### Method 1. Microbiological integrity using *Brevundimonas diminuta:* Partial immersion testing

All testing was performed as described in the NHS YCD – Method 1 sub section 1.4 for partial immersion and according to device IFU.^13,18-19^ Disposable BD syringes (1, 20 and 50 mL) were connected to Chemfort ™ SALs (20 units for each syringe size). Single strength tryptic soya broth (TSB) was withdrawn into each syringe and SAL combination unit from a media fill vial (100mL) pre-fitted with Chemfort™ vial adaptor. All SAL septa were punctured three times including first puncture to withdraw the media into the body of the syringe by mating the SAL with the corresponding vial adaptor component. These three punctures of the septa were performed to present an additional challenge to the CSTD component. All of the manipulations required to prepare the test articles including media draw up were performed using aseptic technique in a validated grade A laminar flow cabinet (HLF6B Envair, UK) sited within a grade B clean room. Following draw up of the media the devices were disinfected (two-step process) following Quality Assurance of Aseptic Preparation Services (QAAPS) protocols and incubated for 7-days at 20-25°C followed by 7-days at 30-35°C to confirm the absence of viable bacterial colonies within the TSB containing syringes, prior to testing.^27^ All test articles that were shown to be free of growth were released for testing in the study at the end of the 14-day quarantine period. All test devices were partially submerged into a vessel containing single strength TSB inoculated with sub-cultured *B. diminuta* (ATCC 11568) and incubated for an additional period of 14-days at 30-35°C. The manipulations for integrity testing were performed in an uncontrolled area with the personnel involved wearing; mop caps, sterile gloves, low shedding clean room coats and masks (MicronClean, UK). The benches were cleaned with 70% alcohol (Helapet, UK) and a sporicidal cleaning agent ProChlor (Contec, UK) before, during and after testing, gloves were regularly cleaned with 70% alcohol (Helapet, UK) and the room had restricted access during the study. The devices post testing were cleaned and inspected for evidence of microbial growth as evidenced by turbidity in the container. Positive control devices consisted of two LL syringe of each size in combination with sterile blind hub as the terminal closure device with the hub left partially open. To demonstrate growth promotion capability two test articles that showed no growth of each syringe size were inoculated with TSB that had been contaminated with *B. diminuta*. The inoculated syringes were then incubated for a period of three days at 30-35°C and inspected visually for signs of microbial growth. The acceptance criteria used for assessment of the devices for evidence of microbial growth was that the test articles should remain clear/ non-turbid and free of growth in the media when inspected visually at the end of test. Any devices that appeared turbid when inspected against clear control samples or showed evidence of microbial growth were assessed as having failed.

### Method 2. Physical integrity-dye intrusion testing using methylene blue (MB) 0.4% w/v

The test articles comprising LL syringe at each volume size fitted with Chemfort™ SAL were connected to a Chemfort™ vial adaptor to allow filling to 75% of maximum volume with MilliQ water. Each Chemfort™ SAL septa were punctured in total three times prior to testing. These three punctures were performed to provide an additional physical challenge to the SAL CSTD component and were performed by mating the SAL to the corresponding CSTD vial adaptor component in accordance with IFU.^19^ A partial internal vacuum was then applied to each test article by pulling out the syringe plunger and securing it using a mechanical screw or pin. Chemfort™ SAL plus LL syringe combinations were filled and immediately placed in a suitable screw topped vessel containing a solution of methylene blue (MB) dye for test. All manipulations of the test devices were performed in a non-sterile uncontrolled laboratory area. Each container of test devices represented a single batch. For the smallest syringe size of 1mL all twenty test articles were accommodated with a positive control (n=1) in one batch. For the largest syringe size of 50 mL five batches were required due to the lower occupancy of the container and hence five positive control tests were performed, one for each batch. For the intermediate size 20mL syringes three batches were required and hence three positive control tests were performed in total. The test articles and positive control devices were submerged in the dye solution and rotated for a total of 2 hours at 45 rpm on a roller mixer.

Positive control devices (n=1) were included in each batch of testing for all syringe volume sizes and comprised Chemfort™ SAL / LL syringe combinations in which a single strand of stainless-steel wire (OD 0.4mm) was inserted, running parallel to the barrel between the plunger seal and the inner barrel wall. The presence of the wire introducing a route of access to the internal compartment of the control device. Different total numbers of positive control syringes for the three sizes were used. This was due to the maximum occupancy for each size combination of BD LL syringe within the cylindrical container. One positive control was included per batch of test articles during testing to verify system suitability of the system. Therefore, for the smallest 1mL syringe tested in combination with Chemfort™ SAL only one positive control device was necessary (n=1). At 50mL which was the largest syringe size tested five batches of test articles were generated and hence five positive control devices (n=5) were required. For the 20mL syringe size 3 batches of test articles were used and hence three positive controls were performed, one per batch (n=3). In total eight positive control tests were performed. Negative controls were performed comprising of all three combinations of SAL/ LL syringe filled with MilliQ water (n=1) that were left at ambient for 2-hours and not immersed in the dye solution or rotated.

At the end of test the Chemfort™ / LL syringe combination devices were removed from the dye solution, washed externally, then inverted twenty times and visually inspected for evidence of dye ingress. In addition, a small volume of each syringe’s contents was removed for quantification using a uv-vis spectrophotometer and validated spectrophotometric method for quantifying the presence of MB dye solution.

A microplate reader set to a detection wavelength of 660 nm was used to read all test solutions in triplicate. Three Quality Control (QC) check standards at low, middle and high concentrations of dye along with MilliQ water (blanks) were read in each plate in triplicate.

The solutions from each syringe combination were analysed in triplicate within the 96-microwell plate. Alongside test solutions, MilliQ water (blank) and the three QC check reference standards (Low, Medium and High) were also analysed in each plate in triplicate. The QC check standards for methylene blue dye were prepared by diluting the 0.4% w/v MB solution in MilliQ water to the following final MB concentrations: 4 × 10^−5^ % w/v (Low); 0.002% w/v (Medium) and 0.4% w/v (High).

For the microplate readings, the average of triplicate MilliQ water blank data (n=3) was subtracted from the average calculated test data (n=3) and the blank corrected results reported for each test device (n=20) comprising the Chemfort™ SAL / LL syringe components.

The dye intrusion test was validated according to ICH guidelines as a limit test with pass or fail outcomes based on the experimentally determined limit of detection (LOD) for detection of colouration of dye visually or when using a Spectrophotometer instrument at 660nm detection wavelength. The pass criteria for visual inspection of the test devices was that the solution inside should remain clear with no detectable blue dye colouration when compared against MilliQ water reference solution and low QC check standard, with the low QC check standard dye solution providing a reference for detectable blue dye colour (1 in 10,000 dilution of 0.4% w/v methylene blue stock solution) at the LOD. The 1:10,000 MB diluted solution was determined at the time of the study to be the LOD as it was lowest amount of dye that could be detected both visually and using a validated spectrophotometer method. All samples from the tested syringe combinations (n=20 for 1 and 50mL syringes, n=21 for 20mL syringes) were visually compared to both MilliQ water (no dye) and to the low concentration QC check standard against a white background. Test devices were assessed as meeting the acceptance criteria for 100% integrity (pass criteria) when in addition to visual inspection spectrophotometric absorbance readings were recorded ≤0.010 (±0.005) mAu at 660nm (<LOD). Where acceptance was not met the result was recorded as a fail indicating positive dye ingress.

## RESULTS

### Microbiological integrity

One of each size of LL syringe were fitted to either a blind hub (n=2) as a positive control (C1, C2, C3) or to a Chemfort™ SAL (n=20) as test article (T1, T2, T3) combinations. Figure 1 below shows the syringe combinations prior to immersion and testing. In figure 1, both test and control syringe combinations were free of microbial growth after draw up and initial incubation immediately prior to test. At the end of the 14-day incubation period at 30-35°C, following the partial-immersion challenge all test combinations were free of microbial growth as can be seen in figure 2 (labelled T1-T3). All positive controls (C1-C3) failed to maintain sterility of the high growth TSB media product at the end of 14-day incubation at 30-35°C as can be seen in figure 2 (labelled C1-C3) which shows clear evidence of growth in the control syringe combinations C1-C3. The data demonstrates the ability of Chemfort™ SAL/ LL combination container systems to maintain 100% integrity under the conditions of the YCD microbiological challenge.^14^

**Figure 1.**
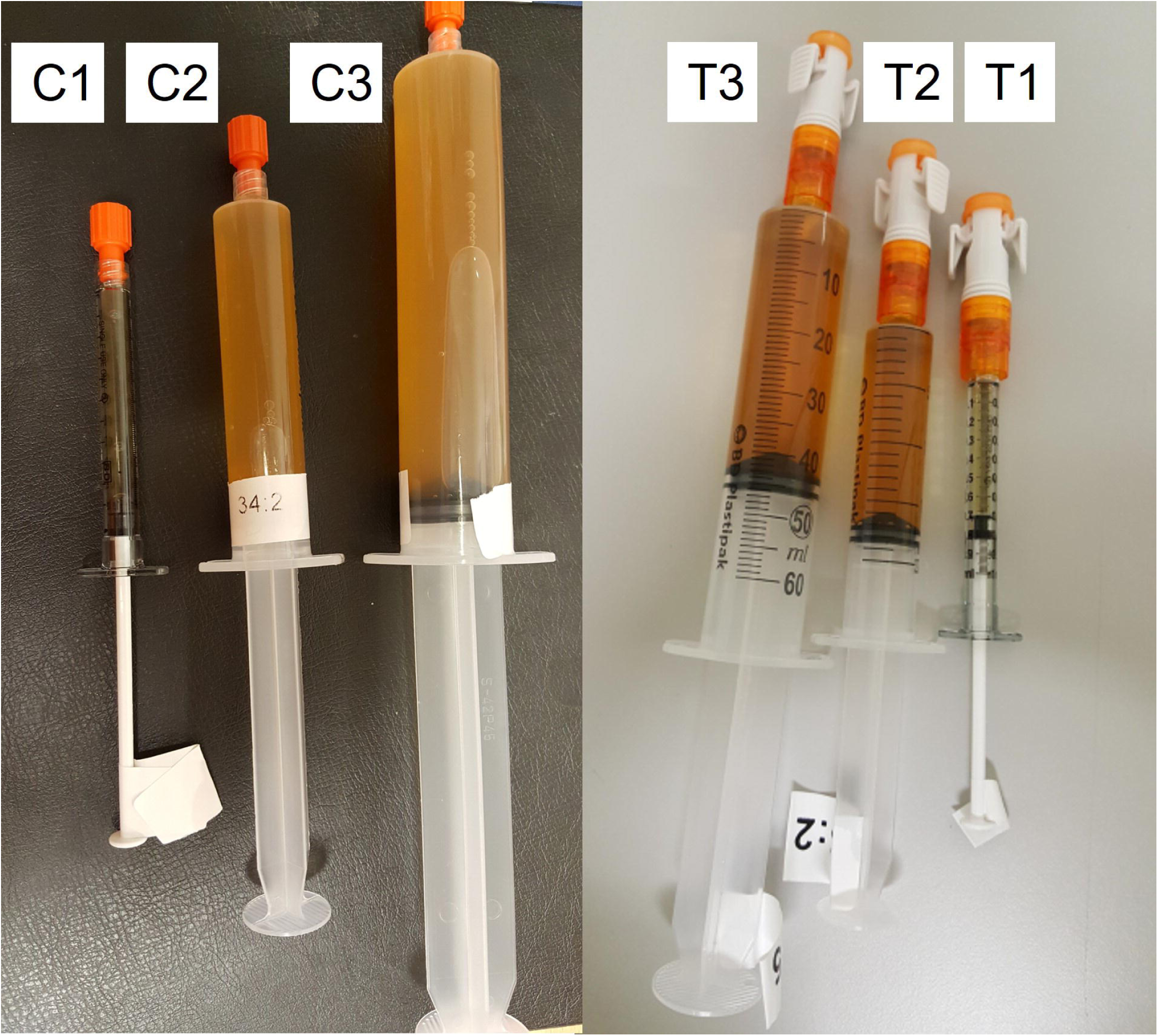
All three volume sizes of test Luer-Locksyringes fitted to either a blind hub as a positive control (C1, C2, C3) or to a Chemfort™ SAL as test article (T1, T2, T3) combinations prior to incubation.

**Figure 2.**
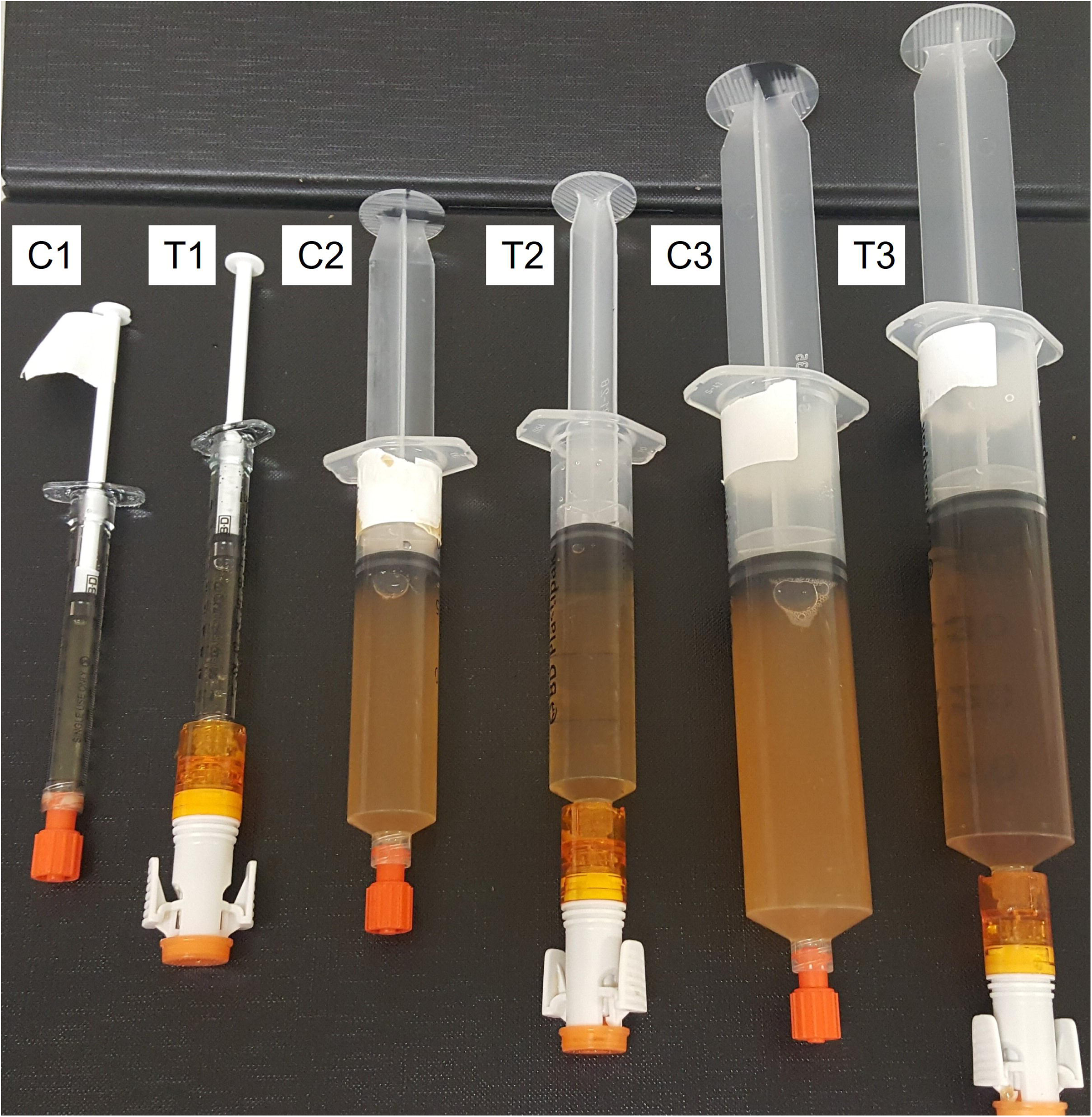
One of each size of LL syringe fitted to either a blind hub as a positive control (C1, C2, C3) or to a Chemfort™ SAL as test article (T1, T2, T3) combinations post 14-day incubation at 30-35°C.

Growth promotion testing demonstrated positive growth of *B. diminuta* in triplicate test devices at each of the three volume syringe sizes tested (n=9) providing evidence of the ability of the growth media to support growth of the challenge organism post testing.

### Physical integrity

Positive controls for all syringe sizes evaluated, tested in combination with SAL resulted in dye intrusion being observed visually and recorded as an absorbance ≥0.010 (±0.005) mAu at 660nm using the spectrophotometer. In every positive control (n=9) a distinct blue colouration was observed visually inside the control syringes. This demonstrated a failure of container integrity and provided evidence for system suitability of the method to detect a positive breach in integrity resulting in dye intrusion.^14^

All negative control articles (n=3) remained free of blue dye at the end of test and hence demonstrated that any blue colouration in the test items was due to MB dye intrusion from immersion. The negative control articles showed absorbance readings ≤0.010 (±0.005) mAu or below LOD at the end of test as determined by a spectrophotometer.

The results from all devices tested and positive controls obtained in this study are presented in Table 1 below for both visual detection and quantitative absorbance measurements at 660nm.

**Table 1.** Summary of the Spectrophotometric data (660 nm) and visual appearance data for test Chemfort™ SAL/ LL syringe combination and control blind hub/ syringe combinations.

| Test Chemfort™ SAL/ LL syringe combination results |  |  |  |  |  | Control blind hub/ LL syringe combination results |  |  |  |  |
| --- | --- | --- | --- | --- | --- | --- | --- | --- | --- | --- |
| Test syringe size | Average absorbance at 660 nm (n=20) | 95% CI | Units tested (n) | Spectrometer Pass/Fail | Visual Pass/Fail | Average absorbance at 660 nm | 95% CI | Units tested (n) | Spectrometer Pass/Fail | Visual Pass/Fail |
| 50mL | -0.004 | 0.001 | 20 | Pass | Pass | 3.352 | 1.094 | 5 | Fail | Fail |
| 20mL | -0.004 | 0.001 | 21 | Pass | Pass | 1.944 | 4.682 | 3 | Fail | Fail |
| 1mL | -0.005 | 0.002 | 20 | Pass | Pass | 0.048 | N/A | 1 | Fail | Fail |

All of the individual 96-microwell plate data for the test Chemfort™ SAL/ LL syringe combination and control blind hub/ LL syringe combinations are presented in the supplemental material section, Table S1 and Table S2.

## DISCUSSION

This study demonstrates that a CSTD syringe adaptor can be used as a terminal closure device for pre-filled syringes containing chemotherapy drug products prepared in aseptic compounding centers. Where advanced preparation and extended storage of the aseptic product is required additional stability data and compatibility data around those aseptic products should also be provided by the device manufacturer to support clinical practice in accordance to the relevant NHS cover document guidance.^13^ The data presented fulfills the requirements of the NHS PQA guidance requirements around sterility data to support the addition of closed system components to pre-filled syringes in aseptic compounding units in the UK and meets the requirements around integrity testing stated in the 2018 NHS guidance document on handling cytotoxics in clinical areas.^13,^ Full integrity of the combination container comprising Chemfort™ SAL/ LL syringe was demonstrated at all three syringe size combinations with no route of access for microbial or physical contamination of the product.

There are three potential routes of entry for external contaminants to access the internal space of the Chemfort™ SAL/ LL syringe combination device: (1) through the luer-to-luer connection between the SAL and commercial LL syringe, (2) through the puncture site of the SAL septa and (3) through the rear of the syringe between the plunger and barrel. In this study routes 1 and 2 were assessed in the microbiological arm of the study and routes 1, 2 and 3 were assessed in the physical arm. The SAL septa were deliberately punctured a total of three times prior to testing, to represent a worst-case scenario. The presence of a puncture site within the septa allows a potential route of access to the internal space of the SAL/ LL syringe combination device. Even after three punctures of the septa of the SAL container closure integrity was demonstrated and found to be 100% in both the microbiological and physical tests.

As the luer-to-luer connection is stringently assessed in both arms of this study the outcomes demonstrate conclusively that the luer-to-luer connection of a 2-piece combination device (CSTD/ LL syringe combination device) is equivalent in device integrity to a syringe device with no luer-to-luer connection present.

No prior published data exists regarding integrity testing of syringe adaptor CSTD components as part of a final container system performed to YCD standards.^13^ However, there are a number of reports from CSTD manufacturers that make claims regarding microbiological integrity testing of CSTD components using different microbial challenge study designs.^28-33^ All report the ability of the respective CSTDs to resist a specific microbial challenge applied to the septa prior to connecting components. The data reported by each author as a microbial ingress test is essentially a microbial assessment for container integrity where the container system comprises CSTD components. In each study microbial contamination of the septa of the vial adaptors is followed immediately by a cleaning disinfection step prior to connection of the CSTD components. As such these microbial challenge studies report the efficacy of the cleaning procedure employed and not the ability of the CSTD components to resist microbial ingress. In the present study a motile organism was employed with the devices immersed over an extended contact time of 14-days at an incubation temperature of 30-35°C allowing optimum growth of the *B. diminuta* challenge organism. This combined with three punctures of the septa membranes prior to immersion represents an extreme challenge for the Chemfort™ SAL/ LL syringe combination container system. Furthermore, the challenge organism length scale is of the order of a few hundred nanometers in diameter and between two and eight microns in length which in combination with its high motility makes it very effective at penetrating breaches in sterile container systems. Finally, it is noteworthy that all of the manufacturer sponsored microbial ingress studies on CSTD components have focused exclusively on the CSTD septa membranes as the main site of entry, whereas the present study assessed overall device integrity from multiple-points of entry not just the CSTD septa.^28-33^

McMichael *et al*. reports microbial integrity testing of PhaSeal (BD) components using media fill vials when accessing the vials over 7 days.^34^ However, all manipulations were performed in an ISO 5 environment where there is not expected to be a microbial challenge to the devices. As such the study reported here is the first study to report an actual microbial challenge to a CSTD component (syringe adaptor lock) when used as part of a terminal container closure system.

In order for a CSTD to meet the NIOSH definition for a closed system transfer device the device needs to demonstrate that: (a) no hazardous drug can escape out from the system, and that (b) no environmental contamination can cross the system boundary.^3^ The authors have previously published data on and demonstrated system performance of Tevadaptor™ CSTD components (Chemfort™ is the second generation of Tevadaptor™) when assessed according to the draft 2016 NIOSH test protocol.^11^ Identical containment information has been proven and is available on file for Chemfort™ (unpublished data). The combined studies fulfil part (a) of the NIOSH CSTD requirement.^3^ The results reported in the present study provides the evidence that Chemfort™ satisfies part (b) of the NIOSH CSTD definition and adds to the body of evidence that demonstrates Chemfort™ being capable of preventing environmental contamination and maintains a sterile barrier when used as part of a final container system.^3^

The data supports the use of Chemfort™ syringe adaptor lock (SAL) to be used as the terminal closure device in combination with Luer-Lock syringes (BD, UK) within aseptic compounding units. Where extended storage of aseptically compounded sterile products used in chemotherapy is required including dose banded products and these are to be prepared using CSTD cap components fitted as terminal closure device the NHS YCD guidance states that additional stability and drug compatibility data is also required from the manufacturer.^13^

Current practice in UK pharmacy technical services involves the use of a sterile blind hub as closure for pre-filled syringes used on aseptic products prepared in advance and stored over extended time periods including chemotherapeutic drugs.^13^ Dose banding of parenteral chemotherapeutic drug products where patient-individualised doses are rounded up or down to predetermined banded doses has been successfully implemented for a number of years in the UK.^35,36^ Banding of chemotherapy doses offer several advantages for the hospital aseptic compounding unit including: reduced patient waiting times, reduction in chemotherapy waiting times, and reduction in drug wastage. However, to leverage economic and patient outcome advantages of advanced aseptic compounding of chemotherapy using dose banding of pre-filled syringes requires: (1) access to extended drug stability data for the drug product, (2) a compounding environment and systems in place for quality control and quality assurance and (3) container integrity data for the final storage device which in the case of a pre-filled syringe is typically a sterile blind hub.^13^ Drug stability and compatibility testing sits outside of the scope of the current study. The present study reports for the first time syringe integrity data for a CSTD component (Chemfort™ SAL) to be used with Luer-Lock syringes (BD) as part of the final container system in pharmacy technical services (PTS) allowing the addition of CSTD components as part of dose banding in the UK.

## CONCLUSIONS

This study reports the testing of Chemfort™ SAL CSTD components when combined with 1mL, 20mL, and 50mL Luer-Lock (BD, UK) syringes in accordance with the NHS yellow cover document (YCD) for syringe integrity and fulfils the requirements of the NHS PQA requirements for addition of CSTD components to pre-filled syringes in UK aseptic compounding units.^13^ All 60 combinations of Chemfort™ SAL device demonstrated 100% integrity across both the microbiological and physical integrity tests. The results support the suitability of Chemfort™ SAL as a terminal closure device for BD LL syringes in aseptic pharmacy technical services. The present study provides the evidence to support Chemfort™ SAL components being added within aseptic services and will be most impactful within compounding centres performing dose banding of chemotherapy drugs where the addition of closed system components as recommended by UK NHS PQA guidelines provide for a safer administration space helping to protect health workers from accidental occupational exposure when handling pre-filled syringes.^3-5,13^

## Data Availability

All data produced in the present study are available upon reasonable request to the authors.

## Acknowledgements and Affiliations

The authors would like to acknowledge and thank Simplivia Healthcare www.Simplivia.com for the provision of a research grant and Chemfort™ medical devices which were used in the testing.

## Ethics approval statement

The authors gained full ethical approval from the supporting organization Biopharma Stability Testing Laboratory ltd of the UK to perform the study.

## Contributorship Statement

Alan-Shaun Wilkinson and Kate Walker were directly involved in the study design, data analysis, and writing of the manuscript. Kate Walker was also involved in planning and data interpretation as well as reporting. Romana Machnikova, Laima Ozolina, Kate Pegg, Navneet Bhogal and Andrew Johnson were all involved in the planning, conduct, acquisition of data, data interpretation, data checking and analysis.

