## Supplemental materials for "Integrity performance assessment of a Closed System Transfer Device syringe adaptor as a terminal closure for Luer-Lock syringes"

**Table S1. Spectrophotometric data set complete recorded on an Epoch microplate reader (Epoch, Biotek UK) for all test sample combinations of Chemfort™ SAL/ LL syringes at a detection wavelength of 660 nm.**

| Chemfort™ SAL/ LL 50mL test syringes | | | Chemfort™ SAL/ LL 20mL test syringes | | | Chemfort™ SAL/ LL 1mL test syringes | | |
|---|---|---|---|---|---|---|---|---|
| **Sample** | **Average abs. at 660 nm** | **Pass/Fail** | **Sample** | **Average abs. at 660 nm** | **Pass/Fail** | **Sample** | **Average abs. at 660 nm** | **Pass/Fail** |
| Syringe 1 | -0.004 | Pass | Syringe 1 | -0.002 | Pass | Syringe 1 | -0.005 | Pass |
| Syringe 2 | -0.003 | Pass | Syringe 2 | -0.004 | Pass | Syringe 2 | -0.004 | Pass |
| Syringe 3 | -0.003 | Pass | Syringe 3 | -0.004 | Pass | Syringe 3 | -0.002 | Pass |
| Syringe 4 | -0.003 | Pass | Syringe 4 | -0.003 | Pass | Syringe 4 | -0.004 | Pass |
| Syringe 5 | -0.003 | Pass | Syringe 5 | 0.001 | Pass | Syringe 5 | -0.007 | Pass |
| Syringe 6 | -0.004 | Pass | Syringe 6 | -0.002 | Pass | Syringe 6 | -0.006 | Pass |
| Syringe 7 | -0.002 | Pass | Syringe 7 | -0.002 | Pass | Syringe 7 | -0.003 | Pass |
| Syringe 8 | -0.005 | Pass | Syringe 8 | -0.003 | Pass | Syringe 8 | -0.009 | Pass |
| Syringe 9 | -0.004 | Pass | Syringe 9 | -0.005 | Pass | Syringe 9 | -0.003 | Pass |
| Syringe 10 | -0.004 | Pass | Syringe 10 | -0.004 | Pass | Syringe 10 | -0.004 | Pass |
| Syringe 11 | -0.007 | Pass | Syringe 11 | -0.005 | Pass | Syringe 11 | -0.008 | Pass |
| Syringe 12 | -0.006 | Pass | Syringe 12 | -0.006 | Pass | Syringe 12 | -0.008 | Pass |
| Syringe 13 | -0.005 | Pass | Syringe 13 | -0.003 | Pass | Syringe 13 | -0.007 | Pass |
| Syringe 14 | -0.007 | Pass | Syringe 14 | -0.006 | Pass | Syringe 14 | -0.006 | Pass |
| Syringe 15 | -0.005 | Pass | Syringe 15 | -0.005 | Pass | Syringe 15 | -0.004 | Pass |
| Syringe 16 | -0.004 | Pass | Syringe 16 | -0.004 | Pass | Syringe 16 | -0.008 | Pass |
| Syringe 17 | -0.004 | Pass | Syringe 17 | -0.004 | Pass | Syringe 17 | -0.005 | Pass |
| Syringe 18 | -0.004 | Pass | Syringe 18 | -0.004 | Pass | Syringe 18 | 0.006 | Pass |
| Syringe 19 | -0.009 | Pass | Syringe 19 | -0.010 | Pass | Syringe 19 | -0.010 | Pass |
| Syringe 20 | 0.001 | Pass | Syringe 20 | -0.008 | Pass | Syringe 20 | -0.005 | Pass |
| | | | Syringe 21 | -0.006 | Pass | | | |

**Table S2. Spectrophotometric data set complete recorded on an Epoch microplate reader (Epoch, Biotek UK) for all combination devices of sterile blind hub/ LL syringes as positive control samples with detection at wavelength of 660 nm.**

| Sterile blind hub/ LL 50mL positive control syringe combinations | | Sterile blind hub/ LL 20mL positive control syringe combinations | | Sterile blind hub/ LL 1mL positive control syringe combination | |
|---|---|---|---|---|---|
| **Sample** | **Average absorbance at 660 nm** | **Sample** | **Average absorbance at 660 nm** | **Sample** | **Average absorbance at 660 nm** |
| P1 | 4.032 | P1 | 1.115 | P1 | 0.048 |
| P2 | 3.655 | P2 | 4.101 | | |
| P3 | 3.895 | P3 | 0.615 | | |
| P4 | 3.329 | | | | |
| P5 | 1.850 | | | | |

### Validation of the Spectrophotometric method:

The methylene blue dye intrusion test method was validated as a limit test according to ICH guidelines. That is to say it results in either a pass or a fail outcome. The limit of detection (LOD) for dye penetration should according to ICH guidelines correspond to the smallest level of dye added to a product that is still consistently detectable. Validation of the spectrophotometric method was performed in accordance with NHS PQA guidance for Pharmaceutical Quality Control Analytical Methods and ICH Q2 (R1) guidance for validation of analytical methods.[1,2]

For specificity testing in accordance with ICH guidelines the dye intrusion method was assessed as to whether it demonstrated that dye penetration resulted in the expected coloration visually and/or spectroscopically and that dye was detectable for a given drug product formulation. When test articles were filled with MilliQ water alone no colouration or absorbance at 660nm was detected. When the same test articles were then filled with the methylene blue dye at a concentration of ~1.2 microMolar a blue dye coloration was observed and an absorbance at 660nm was detected with circa ~0.010 Absorbance units complying with the ICH guidelines for specificity.

Detector response linearity was demonstrated from 0.78 to 12.5 microMoles per Litre methylene blue (MB) in MilliQ water with $R^2$ values (correlation coefficient) of 0.997, 0.993, 0.993, 0.993, 0.995, 0.997 (n=6) with two operators. Slopes from calibration curves were determined at 0.0094, 0.0097, 0.01, 0.0091, 0.0098 and 0.0098 without forcing data through the origin. All linearities were performed by the same two operators over five days. Interday precision and accuracy were performed over five days (n=6) with replicate readings (n=3) at a concentration of 6.25 microMolar MB. The mean absorbance at 660nm for the precision standard at 62.5 microMolar concentration was 0.056±0.002 mAb (±StdDev) with relative standard deviations (RSDs) not greater than 3.5% at this concentration level. An example of a linearity performed is provided below in figure 1S below.

File: Supplementary materials

Figure 1S. Example linear plot of methylene blue (MB) dye absorbance at 660nm versus concentration of methylene blue (MB) dye in MilliQ water (microMolar).

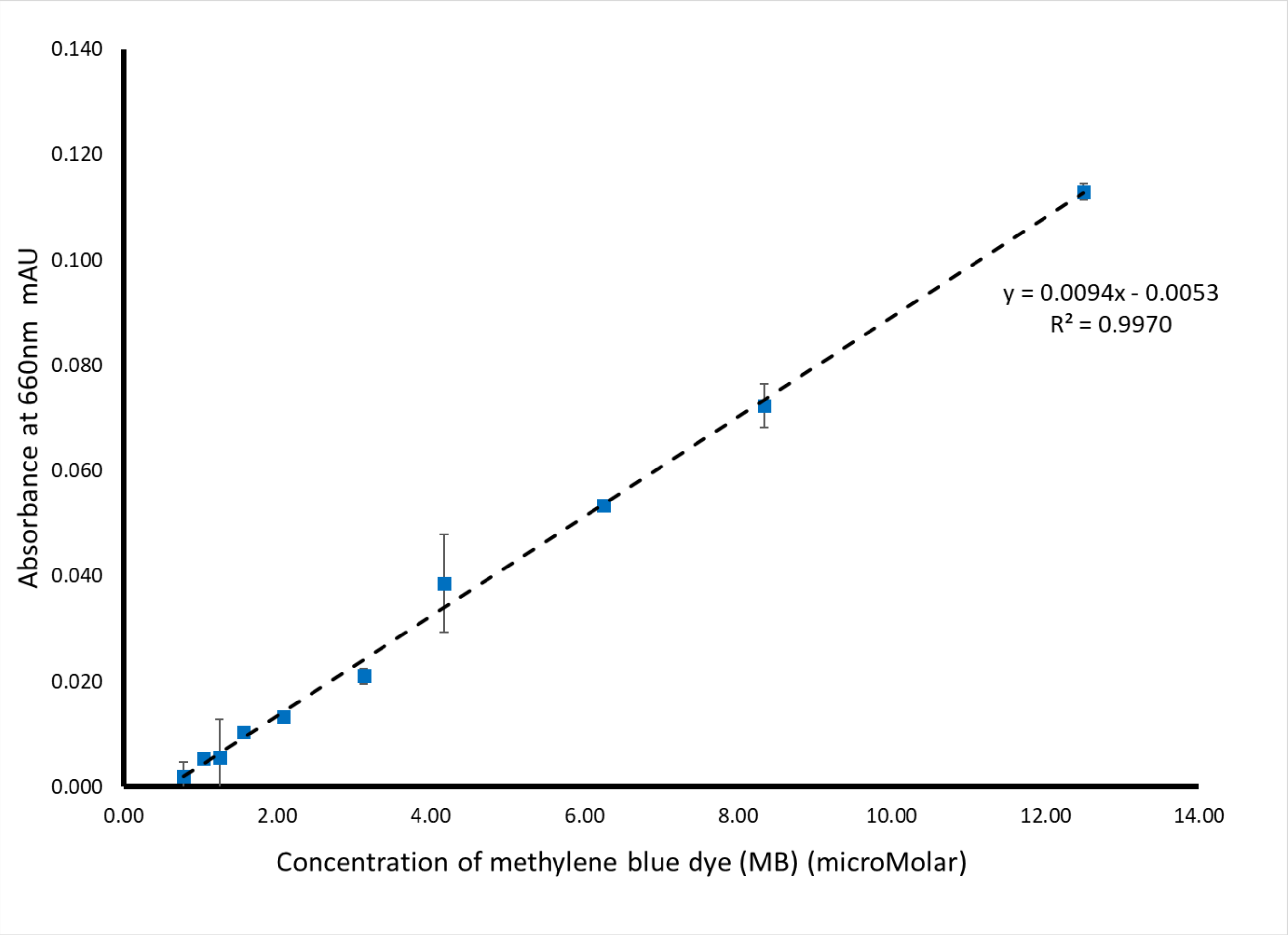


The limit of detection (LOD) was calculated in accordance with ICH Q2 (R1) section 6.3 using equation 1 below and limit of quantitation was calculated using equation 2 below.

Equation 1: LOD = (3.3) *(A)/(B)

Equation 2: LOQ = (10) *(A)/(B)

Where A is the standard deviation of intercept for n=6 linearity determinations and numerically equal to 0.01.

Where B is the slope of the linear correlation and has a numerical value of 0.010 mAu micro Moles$^{-1}$ Litres.

The average LOD over six validation runs was calculated to be 0.43 microMolar (equivalent to a dilution of 1:30,000 of the working MB solution) and the average LOQ over the same six validation runs was calculated to be 1.29 microMolar (equivalent to a dilution of 1:10,000 of the working MB solution.

File: Supplementary materials

#### References

1. Guidance on the Validation of Pharmaceutical Quality Control Analytical Methods - NHS Pharmaceutical Quality Assurance Committee March 2005. www.medicinesresources.nhs.uk/en/Communities/NHS/UKQAInfoZone

2. ICH Q2(R1) Validation of Analytical Procedures: Methodology www.ich.org/products/guidelines.html
